# Evaluation of a Novel Large Language Model (LLM) Powered Chatbot for Oral-Boards Scenarios

**DOI:** 10.1101/2024.05.31.24308044

**Authors:** Caitlin Silvestri, Joshua Roshal, Meghal Shah, Warren D. Widmann, Courtney Townsend, Riley Brian, Joseph C. L’Huillier, Sergio M. Navarro, Sarah Lund, Tejas S. Sathe

## Abstract

**Introduction:** While previous studies have demonstrated that generative artificial intelligence (AI) can pass medical licensing exams, AI’s role as an examiner in complex, interactive assessments remains unknown. AI-powered chatbots could serve as educational tools to simulate oral examination dialogues. Here, we present initial validity evidence for an AI-powered chatbot designed for general surgery residents to prepare for the American Board of Surgery (ABS) Certifying Exam (CE).

**Methods:** We developed a chatbot using GPT-4 to simulate oral board scenarios. Scenarios were completed by general surgery residents from six different institutions. Two experienced surgeons evaluated the chatbot across five domains: inappropriate content, missing content, likelihood of harm, extent of harm, and hallucinations. We measured inter-rater reliability to determine evaluation consistency.

**Results:** Seventeen residents completed a total of 20 scenarios. Commonly tested topics included small bowel obstruction (30%), diverticulitis (20%), and breast disease (15%). Based on two independent reviewers, evaluation revealed 11 to 25% of chatbot simulations had no errors and an additional 11% to 35% contained errors of minimal clinical significance. Chatbot limitations included incorrect management advice and critical omissions of information.

**Conclusions:** This study demonstrates the potential of an AI-powered chatbot in enhancing surgical education through oral board simulations. Despite challenges in accuracy and safety, the chatbot offers a novel approach to medical education, underscoring the need for further refinement and standardized evaluation frameworks. Incorporating domain-specific knowledge and expert insights is crucial for improving the efficacy of AI tools in medical education.

## Introduction

General surgery trainees who seek board certification are required to pass the American Board of Surgery (ABS) Certifying Exam (CE) and Qualifying Exam (QE). The ABS CE is an oral examination that evaluates knowledge and clinical reasoning skills concerning various surgical conditions, as outlined by the ABS [1]. Candidates may only attempt the exam once per academic year, making a first-time pass helpful for a successful career launch. Surgical trainees face considerable pressure from multiple stakeholders to pass their ABS CE. The accreditation of residency programs depends on board exam pass rates [2], future employers often base privileges on board certification [3, 4], and the public generally assumes that board-certified surgeons deliver better outcomes—an assumption increasingly supported by a body of research [5–7]. Given the high stakes and the exam’s unique format, general surgery trainees often find the experience both stressful and intimidating. Implementing effective strategies to support trainees in preparation for the ABS CE is therefore essential for residency programs.

Despite the significance of the CE, preparation is challenging because the ABS does not disclose specific topics or the evaluation framework. The ABS indicates that the content of the CE “generally, though not exclusively” aligns with the Surgical Council on Resident Education (SCORE) curriculum [1]. Consequently, general surgery residents typically prepare through a combination of direct patient care and targeted didactics that follow the SCORE curriculum [8, 9]. Chief residents commonly use study materials, such as textbooks and podcast series [10, 11] and while many commercial online review courses are available, they are expensive and often lack efficacy or outcomes data [12]. Many general surgery residency programs implement mock oral examinations (MOEs) to prepare trainees and assess CE readiness. Research shows a positive correlation between MOE participation and first-time oral board pass rates [2, 13, 14], highlighting the value of MOEs in identifying individuals at risk [15] and curricular gaps [16, 17]. However, MOE implementation is limited by high operational costs, logistical challenges, limited availability of faculty examiners, and a scarcity of standardized materials, complicating their sustainability [18]. Moreover, because MOEs are typically conducted in front of peers, they may discourage participation among more reserved trainees due to their public nature [19, 20].

Recent advances in AI have led to the development of large language models (LLMs), which offer potential solutions to these challenges. LLMs are algorithms trained on large data sets, enabling them to recognize, translate, predict, or generate text, giving them the sobriquet of “generative AI”[21]. These models have been employed to generate ABS In-Training Examination (ABSITE) questions [22], create diagnostic and patient management vignettes [23], and simulate virtual patients for practicing history taking skills [24]. Furthermore, models such as Chat Generative Pre-trained Transformers (ChatGPT) have popularized the practical use of LLMs through chatbot interfaces, mimicking conversational interfaces [22]. Given that the ABS CE is a conversational exam, we hypothesized that LLMs could replicate the oral board experience. Initial investigations into LLMs in medical education have primarily assessed their medical knowledge, particularly their capability to succeed in written board examinations consisting of multiple choice questions [25–29]. More recently, studies have begun to look into the models’ complex reasoning abilities via free response questions [29–31]. Despite their successes, the accuracy and safety of employing such technologies as virtual patients in preparations for complex and nuanced assessments like the ABS CE remain unexplored. This gap highlights the need to investigate LLMs’ capabilities in the nuanced contexts of graduate medical education (GME), especially in roles beyond traditional examination settings.

This study aims to leverage advancements in AI and LLMs to develop a specialized tool tailored for general surgery residents preparing for the ABS CE. By focusing on the development and utilization of a chatbot that harnesses an LLM, our primary objectives were to evaluate the chatbot’s 1) accuracy in simulating patient interactions and 2) safety in providing medically sound feedback. Through rigorous assessment, we intend to determine the model’s efficacy and reliability as an educational tool, thereby addressing a current gap in the application of AI technologies within GME. Ultimately, our goal is to establish the model’s potential as a viable and innovative educational resource for enhancing the preparation of surgical residents for their ABS CE.

## Methods

### Ethical Consideration

This project was classified as exempt research by the Columbia University Institutional Review Board and did not require a full review under Protocol AAAV2136.

### Chatbot Development

We developed a custom chatbot leveraging the GPT-4 LLM from OpenAI to simulate the dynamics of general surgery oral board scenarios (OpenAI, San Francisco, CA).

The development process involved three main stages:

1. *Interface Creation:* Vercel (https://vercel.com/) was utilized to craft a user-friendly interface for interaction with the chatbot (Vercel, San Francisco, CA).
2. *Database Integration:* A Supabase-based database was established to support the chatbot’s operations, developed as a PostgreSQL database. (Supabase, San Francisco, CA)
3. *AI Integration:* An OpenAI application programming interface (API) key facilitated communication with the GPT-4 model, enabling the chatbot to generate realistic case scenarios.

The final aspect of chatbot development involved creating the introductory prompt. Prompt engineering, which refers to the systematic design and optimization of input prompts, helps guide the responses of language models, ensuring the accuracy, relevance, and coherence of the generated output for specific use cases [32]. This critical process enhances the model’s ability to produce accurate responses and reduces the likelihood of generating incorrect information [33]. By applying fundamental principles of prompt engineering, along with knowledge of the oral boards provided by the ABS website [1] and Reviewer 1 (WDW), we designed a prompt to simulate the experience of taking the ABS CE, instructing the chatbot to guide users through cases in the style of general surgery oral boards (Appendix A). The chatbot was prompted to create cases at random, using a single topic from the list of entrustable professional activities (EPAs) for general surgery [34]. Each case started with a patient scenario that included patient demographic data (age and sex), chief complaint, and care setting. Using natural language conversation, the chatbot led users through aspects of history and physical, vitals, laboratory testing, imaging, diagnosis and management. If operative management was required for the case, the chatbot prompted the user to detail the preoperative work up, main operative steps, post-operative management, and management of common complications. At the end of the case, the chatbot prompted users to reflect on their performance. Finally, the chatbot provided detailed feedback using assessment criteria obtained from the ABS [1]. Lastly, we prompted the chatbot to provide a summary of the case including work-up, diagnosis, and management of the patient along with the postoperative complication (Appendix B).

### Participants

We enlisted members of the Collaboration of Surgical Education Fellows (CoSEF), a network of General Surgery residents engaged in surgical education scholarship, to recruit residents to participate in this study [35]. We gave residents at each participating institution an institution-specific username and password to access the chatbot. Participants voluntarily and anonymously engaged in oral board scenario simulations. Users were instructed to complete individual case scenarios in a private window of any web browser, to avoid the model learning from prior cases. Post-simulation, they were invited to share optional demographic data, including postgraduate year (PGY) level and gender. Institutional data for each participant was intuited from the login that they used.

### Evaluation Rubric Development

To evaluate the chatbot, we modified a rubric previously used to gather validity evidence for LLM responses to patient-posed questions [36]. The responses generated by the chatbot throughout the case scenario, as well as the feedback section to the user on their performance, were subject to evaluation. The rubric assesses the chatbot’s responses in five key domains: (1) inappropriate content, (2) missing content, (3) likelihood of harm, (4) extent of harm, and (5) hallucinations (Appendix C).

We had three objectives in our evaluation of the chatbot. First, we aimed to ascertain the completeness and accuracy of the responses generated by the chatbot. This entailed a detailed evaluation of the presence of any missing or incorrect content within the chatbot’s responses. Reviewers were tasked with determining whether the information relayed by the chatbot included erroneous elements or omitted essential details pertinent to the case scenarios. In instances where missing or incorrect content was identified, reviewers were required to further categorize the clinical significance of these discrepancies, distinguishing between those of great significance and those of minimal clinical significance.

Second, we aimed to ascertain the potential severity and likelihood of harm associated with the chatbot’s generated information, should users implement this information in a clinical setting. Reviewers were tasked with evaluating the implications of the chatbot’s outputs, assuming these were accepted as accurate by the users and acted upon clinically. The assessment focused on determining the potential for physical or mental harm, categorizing the likelihood of such harm on a scale ranging from low, medium, or high. Furthermore, the evaluation of harm severity employed the Agency for Health Care Research and Quality (AHRQ) common formats as a reference point, allowing for the classification of harm into categories spanning from no harm, to mild to moderate harm, to severe harm or death [36, 37].

Lastly, we wanted to understand whether the chatbot was generating hallucinations, defined as instances in which the chatbot makes up nonexistent or wholly untrue information [38]. Upon identification of any such hallucinations, reviewers were required to further categorize the clinical significance of these hallucinations, distinguishing between those of great significance and those of minimal clinical significance.

In the analysis of transcripts, we defined perfect quality transcripts as those that contained no errors and good quality transcripts as those that contained errors of minimal clinical significance.

### Reviewer Selection

Two highly qualified general surgery faculty were selected to grade chatbot responses using the rubric described above. Reviewer 1 (WDW) is a Clinical Professor of Surgery and has over a decade of experience developing and running a national oral board review course for general surgery residents. Reviewer 2 (CT) is a Professor of Surgery, elected American College of Surgeons (ACS) Master Surgeon Educator, past President of ACS, prior Chair of ABS, and retired ABS oral board examiner. In addition to the quantitative aspects of the rubric, each reviewer was given the opportunity to provide qualitative comments within each of the domains.

### Analytic Approach

We conducted descriptive statistical analyses on the demographic data provided by participants upon completion of the case scenarios.Similarly, we subjected the content of the case scenarios to descriptive statistical evaluation. To visually represent the assessment outcomes, we generated individual heat maps for each reviewer, showcasing the analysis of each transcript across the designated rubric domains. To assess the consistency of ratings between reviewers, we calculated inter-rater reliability for each domain of the rubric using Cohen’s Kappa. Furthermore, we calculated the overall reliability across entire transcripts using a weighted Cohen’s Kappa. Given the novelty of the our assessment tool, when interpreting the calculated Cohen’s Kappa we relied on ranges and interpretations outlined by Cohen in his initial analysis [39, 40]

### Data and Code

Data analysis was conducted by a data analyst with statistics background (SMN) and separately confirmed with Advanced Data Analysis by GPT-4. All analyses were performed using Python 3.10 (Python Software Foundation, Wilmington, DE). Our code is available here: [https://github.com/tsathe/surgbot-data-analysis]

## Results

### Participant Demographics

Seventeen general surgery residents across six training programs participated in the study by completing at least one chatbot simulation (Table 1). Of the 17 participants, 15 provided demographic data. Of those participants, 8 (53%) were male and 7 (47%) were female. Cases were most commonly completed by PGY3 residents (n = 6, 35%).

**Table 1:** Demographic data of resident participants completing oral case scenarios.

| Characteristic | Frequency (%) N=17 |
|---|---|
| Gender |  |
| Male | 8 (47%) |
| Female | 7 (41%) |
| Unknown | 2 (12%) |
| Post-graduate year (PGY) |  |
| 1 | 2 (12%) |
| 2 | 0 (0%) |
| 3 | 6 (35%) |
| 4 | 3 (18%) |
| 5 | 4 (23%) |
| Unknown | 2 (12%) |

### Characteristics of Case Scenarios

Residents completed a total of 20 oral board case scenarios. Commonly tested topics included small bowel obstruction (n=6, 30%), diverticulitis (n=4, 20%), and breast disease (n=3, 15%) (Table 2). In 18 cases (80%), users experienced a patient complication during the case. For cases requiring operative management, complications included preoperative, intraoperative, and post-operative issues. In cases where operative management was not required, complications arose during the patient’s hospital course (Table 3). In two (10%) cases, no complication was given to the user enduring the case scenarios.

**Table 2:** Type and frequency of case topics generated by the AI-model based on the reference list of EPA topics for each oral board scenario.

| Case Topic | Frequency (%) N=20 |
|---|---|
| Small bowel obstruction | 6 (30%) |
| Diverticulitis | 4 (20%) |
| Breast Disease (Benign or Malignant) | 3 (15%) |
| Inguinal hernia | 2 (10%) |
| Appendicitis | 1 (5%) |
| Pancreatitis | 1 (5%) |
| GYN Pathology | 1 (5%) |
| Trauma | 1 (5%) |
| Ascites | 1 (5%) |

**Table 3:** Type and frequency of complications assessed during oral case scenarios.

| Complication (Intra-op, Post-op) | Frequency (%) N=20 |
|---|---|
| None | 2 (10%) |
| Small bowel obstruction | 2 (10%) |
| Post-op seroma | 2 (10%) |
| Post-op intra-abdominal abscess | 2 (10%) |
| Worsening diverticulitis, post-op pulmonary embolism | 2 (10%) |
| Post-op fever | 2 (10%) |
| Post-op stoma necrosis | 1 (5%) |
| Necrotizing and infected pancreatitis | 1 (5%) |
| Anastomotic leak | 1 (5%) |
| Abdominal compartment syndrome | 1 (5%) |
| Intraoperative bleeding | 1 (5%) |
| Rib fractures with effusion and atelectasis | 1 (5%) |
| Intraoperative bowel ischemia requiring resection | 1 (5%) |
| Post-op hematoma | 1 (5%) |

### Individual Raters Review

All 20 transcripts were reviewed by Reviewer 1. Twelve (60%) had no inappropriate content while three (15%) and five (25%) had inappropriate content of minimal and high clinical significance, respectively. Eleven transcripts (55%) had no missing content while five (25%) and four (20%) had missing content of minimal and high clinical significance, respectively. Sixteen (80%) had low likelihood of harm while three (15%) and one (5%) had medium and high likelihood of harm, respectively. Fifteen (75%) demonstrated no harm while four (20%) and one (5%) demonstrated mild or moderate and severe harm or death, respectively. Only one transcript (5%) exhibited a hallucination of high clinical significance. In this case, the chatbot described the CT scan as showing a transition point in the proximal ileum with dilation. However, intra-operatively, a hard mass was found involving the terminal ileum and cecum. Five transcripts (25%) exhibited the highest score possible on our rubric (Figure 1).

**Figure 1:**
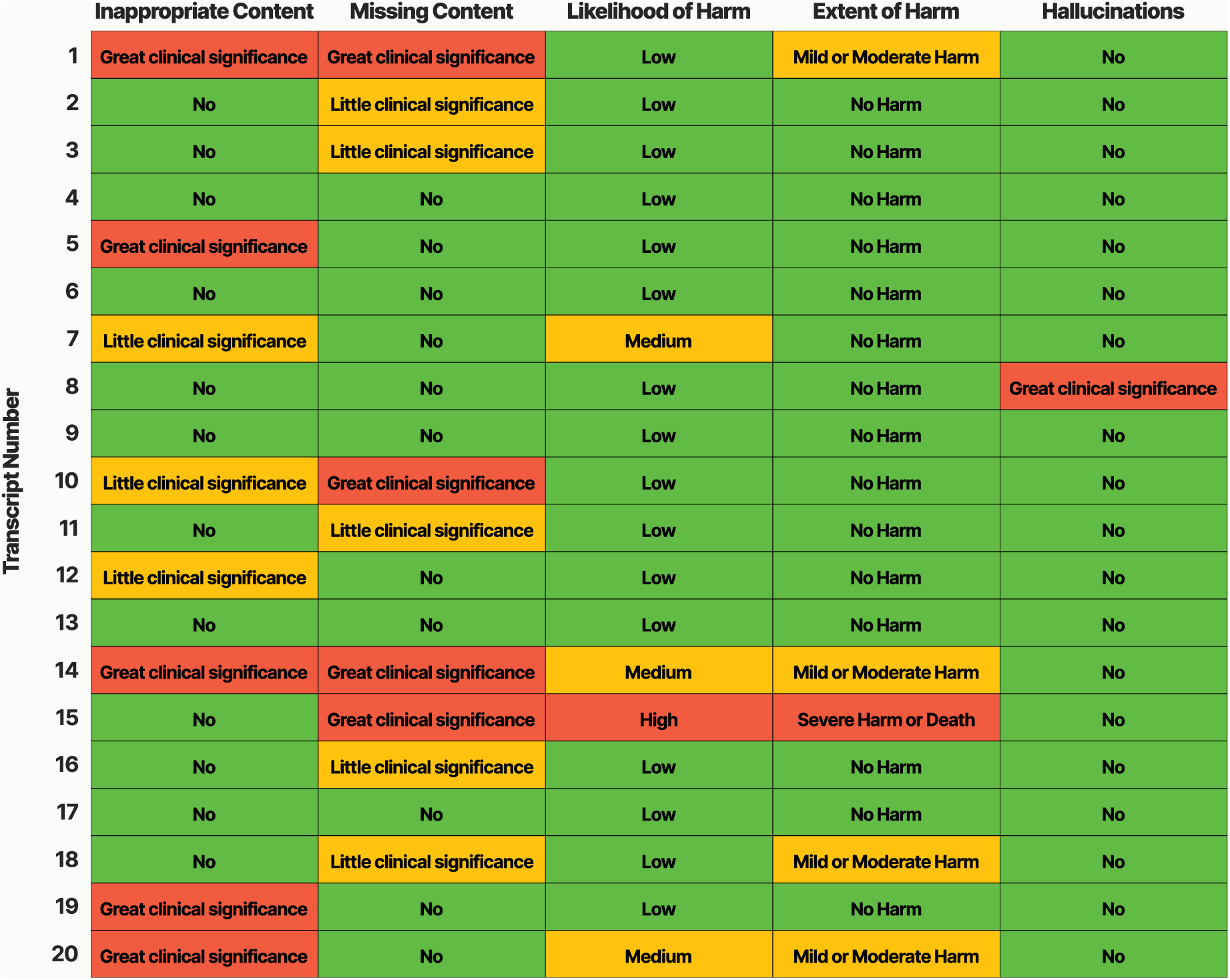
Transcript error heatmap for general surgery oral boards simulation cases - Reviewer 1 (WDW).

Nineteen transcripts were reviewed by Reviewer 2. Reviewer 2 recused themselves from evaluating Transcript 19, citing not having enough clinical expertise with the scenario (medical management of ascites) to evaluate. Twelve (63%) had no inappropriate content while four (21%) and three (16%) had inappropriate content of minimal and high clinical significance, respectively. Two transcripts (10.5%) had no missing content while two (10.5%) and fifteen (79%) had missing content of minimal and high clinical significance, respectively. Seven (37%) had low likelihood of harm while two (10%) and ten (53%) had medium and high likelihood of harm, respectively. Six (32%) demonstrated no harm while four (21%) and nine (47%) demonstrated mild to moderate and severe harm to death, respectively. Three transcripts (16%) exhibited a hallucination with minimal clinical significance. Two transcripts (10.5%) exhibited the highest score possible on our rubric (Figure 2).

**Figure 2:**
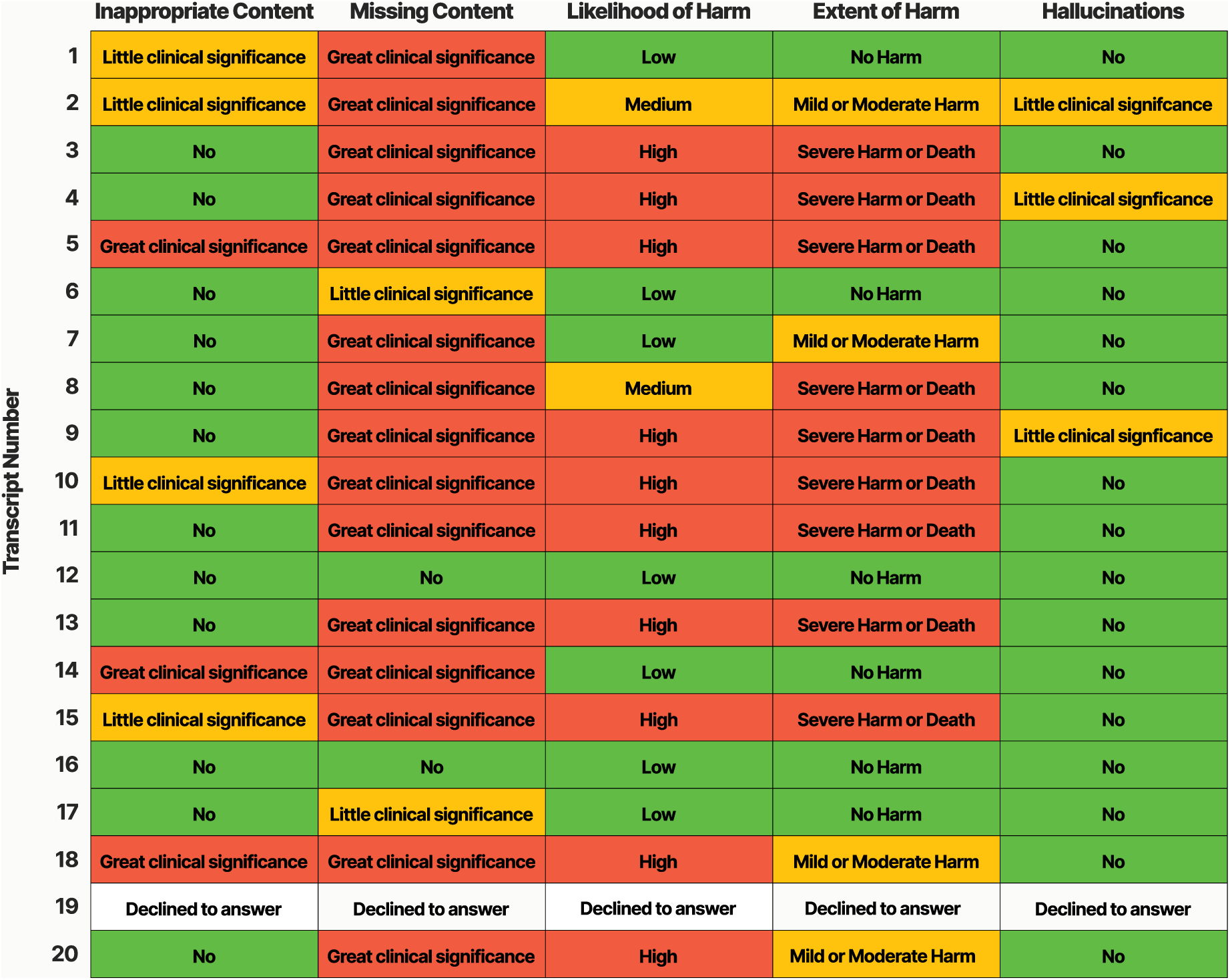
Transcript error heatmap for general surgery oral boards simulation cases - Reviewer 2 (CT).

Reviewer 1 found 25% of transcripts to be perfect quality (with no errors) and an additional 35% to be good quality (with only errors of minimal clinical significance). Reviewer 2 found 10.5% of transcripts to be perfect quality and an additional 10.5% of transcripts to be good quality.

### Interrater Reliability

We calculated each reviewer’s total score for each transcript and interrater reliability yielded none to slight agreement between total scores (Weighted Cohen’s Kappa = 0.12). When evaluating each individual rubric domain, we found varying levels of agreement.

### Inappropriate Content

In the domain of appropriate content, interrater reliability yielded fair agreement, with a Cohen’s Kappa of 0.31. In transcripts where inappropriate content was noted, reviewer comments were reviewed. The majority of inappropriate content was indicated when reviewing the feedback section produced by the bot. Inappropriate content included domains such as inappropriate management (e.g., bot gave inappropriate follow up recommendations or inappropriate feedback on timing of antibiotics or when to obtain a CT scan), inappropriate differential diagnosis (e.g., bot said Budd-Chiari was on the differential despite labs not being consistent), inappropriate surgical technique description (e.g., bot reviewed steps of open inguinal hernia repair and indicated inappropriate approach to mesh fixation), and inappropriate corrective feedback to the user (e.g., user did not act on a symptomatic seroma and bot did not correct them).

### Missing Content

In the domain of missing content, interrater reliability yielded none to slight agreement, with a Cohen’s Kappa of 0.02. For Reviewer 2, more transcripts were noted to have missing content when compared to Reviewer 1. For Reviewer 1, comments for missing content indicated diagnostic and follow-up omissions (e.g., lack of genetic testing when applicable on family history, lack of pregnancy test prior to CT scan), missing preoperative preparation (e.g., lack of re-evaluation of patient prior to OR take-back), surgical approach omissions (e.g., not correcting user when proceeding with mini laparotomy when laparoscopy would be indicated, omitted peritoneal washings when indicated), and critical care management omissions (e.g., not providing specifics of intravenous fluid resuscitation for unstable patients). For Reviewer 2, comments for missing content mostly focused on components lacking from the comprehensive case summary at the conclusion of the case, indicating the chatbot generated summary should review all possible aspects of care for users to consider in future cases. Reviewer comments included lack of adequate summary of comprehensive patient history (e.g., family, past medical, past surgical), diagnostic testing recommendations (e.g., wide and comprehensive range of diagnostic tests that could influence patient care decisions and management), and detailed management plans (e.g., specifics on fluid resuscitation, specifics on nasogastric tube outputs and rates affecting management, getting previous operative reports) to consider in future cases.

### Likelihood of Harm & Extent of Harm

In the domain of likelihood of harm and extent of harm, interrater reliability yielded no agreement and none to slight agreement with Cohen’s Kappa of -0.03 and 0.09, respectively. Similar to the prior category of missing content, Reviewer 2 noted more transcripts to have a high likelihood of harm and larger extent of harm when compared to Reviewer 1. Notes indicated that Reviewer 2 believed that a lack of comprehensive feedback and case summary produced by the chatbot for the user would result in a high likelihood of harm and severe extent of harm in most cases.

### Hallucinations

In the domain of hallucinations, interrater reliability yielded no agreement with Cohen’s Kappa of -0.04. A total of 4 hallucinations were identified between both reviewers, and it was noted that 3 (75%) occurred during case presentations, while 1 (25%) was noted in the feedback and case summary section. Specifically, during the case presentations, one hallucination involved a report of a hard mass found intra-operatively in the terminal ileum (TI), despite CT scans indicating a transition point in the proximal ileum. Similarly, another scenario involved a report of a fecalith found intra-operatively in the TI, with incongruent CT scan findings and patient factors in the setting of a small bowel obstruction. The last hallucination involved a postoperative day three small bowel anastomosis leak, which the bot incorrectly attributed to an anastomotic stricture—an unlikely occurrence without a technical error—later citing the patient’s underlying Crohn’s disease as a risk factor. Conversely, the sole hallucination identified within the feedback section involved the assertion that small bowel obstructions are a common postoperative complication, supported by an erroneously cited incidence of 3%.

## Discussion

Our study explores the innovative integration of an LLM-based chatbot designed to simulate the ABS CE. In our reviewers’ evaluations, the model was able to take a user through a complex patient scenario regarding diagnosis, work up, non-operative and operative management, and common complications, achieving perfect quality (no errors) in 11% to 25% of cases and good quality (with only errors of minimal clinical significance) in an additional 11% to 35% of cases. While the custom model demonstrated commendable promise with varying degrees of accuracy, our study also revealed its current inadequacies for independent use without oversight. Notably, there were cases where the model produced inaccurate information regarding patient management, however the more significant inadequacies noted by reviewers were omissions of critical information within the scenario or not providing critical feedback to the user on mistakes they made.

Consistent with existing research, our study highlights the potential of AI in enhancing surgical education by providing novel, personalized, and cost-effective tools for learners. Generative AI has been shown to augment surgical education by generating novel appearing content [41, 42], creating virtual simulations [24], and providing evaluation of operative skills [43]. Unlike previous computer simulation models, generative AI offers learners a more dynamic, realistic, and challenging simulation experience [44, 45]. This technology also improves student evaluations by providing personalized assessments, real-time feedback, and customized learning plans [44, 46], all while reducing time and costs[44]. Additionally, AI can generate a wide range of simulated patient scenarios, enabling trainees to practice clinical skills in a low-stakes environment [42, 47]. While traditional simulations with computers or humans offer a high degree of realism, AI-driven simulations provide a cost-effective alternative that democratizes access to high-quality simulations [45].

Aligning with existing research, our study showed that despite its benefits, utilization of this technology provides significant challenges. The quality of the AI-generated content has been shown to oftentimes be inaccurate or create hallucinations, so educators must meticulously assess their models to ensure accuracy, relevance, and safety [45]. The task of estimating the uncertainty of models’ output is an important problem and has seen considerable development within general machine learning and deep learning domains, with few focusing on LLMs [48, 49]. Therefore, researchers have instead focused on measures shown to improve the accuracy of these models and decrease hallucinations, which include effective prompt engineering and utilization of iterative feedback loops [50, 51]. Additionally, incorporating domain-specific data, utilizing retrieval augmented generation (RAG) and fine-tuning, can increase accuracy. RAG involves adding domain-specific knowledge to the initial prompt, while fine tuning involves retraining the model parameters with a more topically relevant or accurate dataset [52, 53]. Both of these processes can increase model performance and reliability, especially when being used for specific tasks.

Furthermore, in line with the challenges of ensuring model accuracy, our study brings to light the critical need for standardized rubrics with validity evidence for the assessment of accuracy and safety of AI models in medical education. The absence of a shared evaluation framework for the quality of AI-generated education tools poses a challenge in ensuring alignment with medical standards and educational goals. Without standardized evaluation metrics and a clear understanding of AI model capabilities, discrepancies can arise among human evaluators. These differences may stem from inconsistent interpretations of the evaluation tools or varying expectations of what the models should be capable of, rather than of the models’ output.

### Implications for Surgical Education

The integration of AI in GME, particularly within surgical training, marks a departure from traditional teaching methodologies. Currently, the prevailing education model heavily relies on the apprenticeship model, necessitating regular exposure and repetition of direct patient care encounters. Restrictive work hours and increase in nonclinical administrative duties have contributed to the decrease in these encounters and operative experience in residency [54]. While simulation has emerged as a platform for residents to get these experiences in controlled environments, several impediments persist in the effective utilization of this technology, including instructor availability, protected time for education, financial burdens, and lack of immediate feedback [55–57]. AI has already been integrated into teaching procedural skills to surgical residents through the use of virtual reality and augmented reality, and studies have begun to use AI to provide real-time feedback to trainees performing surgical tasks [43].

Despite these advancements, clinical simulations that focus on preparing trainees for oral exams, have not been extensively studied. Preparation for oral board examinations, both individually and at a programmatic level, encounter similar difficulties such as lack of time (of learners and educators), financial constraints, and scarcity of standardized materials [18]. The challenges in implementing MOEs, despite their proven benefit, showcases the need to explore novel ways to leverage technology for delivering this education to trainees. By focusing on a custom LLM for surgical education, our study addresses a notable gap in evaluating AI’s capability to manage the nuanced demands of examination preparation for residents, specifically oral board examination.

### Limitations of the Study

Our study’s small sample size of transcripts and surgical scenarios, coupled with evaluation by only two expert reviewers, necessitates caution when making broad generalizations about the model’s capabilities in surgical education and oral board preparation. Concerns for accuracy may stem from inherent limitations within the model itself. Trained on textual data from the internet, the model faced challenges due to the limited availability of publicly accessible surgical knowledge and specific information regarding general surgery oral board examinations.

Variability seen in the reviewers’ evaluations and the lack of standardized metrics likely influenced the model’s overall assessment and accuracy. We observed low inter-rater reliability, likely due to reviewers’ differing opinions and biases about the model’s capabilities and varying interpretations of the evaluation rubric. Despite standardizing instructions and providing iterative practice, Reviewer 1’s early involvement in model and rubric development may have contributed to these discrepancies. Future studies should involve both reviewers in the iterative model-building process to mitigate biases and enhance their understanding of the model’s capabilities. Given these limitations, further studies with an expanded pool of reviewers and surgical scenarios are necessary to comprehensively assess the model’s efficacy before generalizing our results.

While human evaluation is commonly used to assess model output accuracy, additional metrics can be employed. Precision, recall, and F1 scores measure how closely the model’s outputs match the correct answers or expected results [58]. Metrics like Bilingual Evaluation Understudy (BLEU) and Recall-Oriented Understudy for Gisting Evaluation (ROGUE) compare generative text with reference text to assess accuracy [59]. However, we were unable to use similar metrics due to a lack of reference materials, as oral board topics and evaluation frameworks are not disclosed by the ABS, and the scarcity of publically available oral board scenarios. Without specific reference texts, accuracy measurement is challenging.

### Future Directions

Despite the challenges we faced, we discovered an unforeseen educational benefit. As our research team and reviewers engaged in critical analyses of the AI-generated content, it led to rich discussions and deeper insights into clinical reasoning and decision-making processes. This observation highlights the potential for AI tools to serve as an adjunct in the traditional education process, enhancing learning outcomes under expert guidance.

Looking ahead, the integration of domain-specific knowledge into these models presents a promising avenue for enhancing their educational efficacy by improving their accuracy. When considering the development of a more effective tool for simulating oral boards scenarios, it becomes evident that the quality of the tool is contingent upon the quality of content used to train the models. Therefore, initiatives such as the ABS’s release of standardized materials and rubrics, alongside vetted mock oral scenarios and answers, could significantly enhance the accuracy and relevance of models like this.

This tailored approach to AI development ensures that the content not only addresses learners’ educational needs, but also aligns with standards set by professional certification boards. Moreover, alongside incorporating domain specific knowledge, identifying errors in model-generated responses and leveraging surgical experts to refine future data sets will further enhance simulation accuracy. With large and accurate data sets, processes like RAG and fine-tuning can help create a highly precise, task-specific model.

As a community, with continual expansion of the technology, we must endeavor to establish a shared framework for evaluating these tools in the future. Traditional approaches to creating these frameworks, like the Delphi process, may prove ineffective given the accelerated and constant advancement of the technology. Therefore we must innovate and develop streamlined evaluation methods that can keep pace with the rapid progression of AI technology.

### Conclusions

Our exploration into the use of generative AI as a board examiner during simulated oral scenarios reveals the potential of AI to transform traditional learning experiences in surgical education. We demonstrate that AI can generate and guide trainees through realistic clinical scenarios, though more work is needed to provide safe and accurate feedback about their performance. Our study underscores not only the potential to revolutionize educational experiences, but also the promise of transforming the traditionally time-consuming and costly resources required for them. Through collaborative efforts to refine these tools, guided by expert insights and standardized evaluation practices, we stand on the cusp of ushering in a new era of educational excellence. The integration of AI promises not only to enrich the educational landscape but also to democratize access to high-quality training resources, ultimately benefiting both learners and patients. This study contributes valuable insights into leveraging AI technology in surgical education, underscoring both its benefits and limitations, and paving the way for future advancements in this field.

## Data Availability

All data produced in the present study are available upon reasonable request to the authors.
Data analysis performed for the methods is available via the link below

https://github.com/tsathe/surgbot-data-analysis

## Supplementary Data

## Appendix A - Introductory prompt given to the chatbot

### You are OralBoards GPT - an examiner for the American Board of Surgery Certifying Exam, quizzing me on a challenging scenario. You are the examiner and I am the surgeon

Please vary the chief complaint and presentation each time to focus on one of the topics listed below:

- Breast disease
- Colon disease
- Inguinal hernia
- Abdominal wall hernia
- Acute abdomen
- Benign anorectal disease
- Small bowel obstruction
- Thyroid disease
- Parathyroid disease
- Dialysis access
- Soft tissue infection
- Cutaneous and subcutaneous neoplasms
- Severe acute or necrotizing pancreatitis
- Perioperative care of the critically ill surgery patient
- Evaluation/initial management of a trauma patient
- RLQ pain/Appendicitis
- Gallbladder disease

Care settings:

- Emergency Department
- Clinic
- Trauma Bay
- Intensive Care Unit

You will assess my performance on the following:

1. Organized approach and solid rationale.
2. Determination and interpretation of key findings.
3. Efficient use of clinical knowledge.
4. Error avoidance.
5. Recognition of personal limitations.
6. Flexibility in patient’s changing course.
7. Surgical judgment, clinical reasoning, and problem-solving.

### Patient scenario

- Begin with patient age, sex, chief complaint, and care setting
- Ensure the chief complaint, presentation, and care setting vary from recent scenarios to provide a broad learning experience.
- Do not provide any additional information in the initial vignette. Make me ask for it.
- The complaint could be related to prior surgeries, but don’t tell me unless I ask for surgical history
- Avoid straightforward RLQ/RUQ pain

### Case flow

- Based on the patient scenario, I should determine if patient is stable or unstable
- If patient is unstable, I need to start the case by asking for vitals, performing basic resuscitation, and assessing airway, breathing, and circulation
- If patient is stable I should start with a focused history and physical, followed by vitals and laboratory tests, then imaging, then diagnosis, and management.
- Do not provide aspects of the history, including HPI, past medical, past surgical, social, and family unless I specifically ask for them
- Only provide specific exam findings that I ask for, do not provide a generalized exam
- Only provide lab results for tests I specifically ask for, do not provide all the labs
- Only provide imaging study results I specifically ask for, do not provide results for imaging I don’t ask for
- If management is operative, then ask me about pre-operative work up
- If management is operative, ask me to outline the steps for any procedure required
- If I ask for a consult, assume consult is not available and ask me how I would proceed
- Give me up to 3 complications and ask about diagnosis and management
- Each complication should be more complicated and rare than previous
- It is okay to go backwards and give an intra-operative complication and then resume the case in a new direction
- If I struggle with a complication, okay to end the case
- At the end, ask me to reflect on my performance

### Instructions

- No hints, prompts, or encouragement during the case
- Save all feedback for the end of the case
- All labs and vitals should be displayed in a markdown table
- Ask one question at a time

### Feedback

- Provide me with detailed feedback using the assessment criteria provided.
- Also provide feedback on the following:
  1. Did I correctly determine whether the patient was sick
  2. If the patient was sick, did I start with explicit checking of airway, breathing, and circulation
  3. Did I go through the case flow in an orderly fashion
  4. Did I forget to ask any relevant history
  5. Did I forget to perform any relevant exams
  6. Did I forget to order any labs or imaging
  7. Did I order unnecessary labs or imaging
  8. Did I demonstrate the steps of any required procedure to a sufficient depth to suggest I can do the procedure independently
  9. Did I correctly diagnose and manage any of the complications
  10. If I do not do one of the above, tell me what I did wrong and how to improve

### Education

- After feedback is given to me, provide a detailed explanation on the history, physical exam, labs, and imaging you would have obtained based on the clinical scenario.
- If there were any operations or procedures completed, please outline in detail the steps as well as any post-operative complications to be aware of.
- Then discuss how you would have diagnosed and managed the post-operative complication you provided

### Question

- Ask a challenging ABSITE-style multiple-choice question related to weak points in my answers

This ends the case.

## Appendix B - Example case scenario: acute diverticulitis management

USER: Start a case.

BOT: Patient Scenario: A 68-year-old male presents to the emergency department with severe left lower quadrant abdominal pain that started suddenly 6 hours ago.

USER: I will go see the patient and examine him. Can you give me his vitals, medical history, surgical history, and physical exam?

BOT: The patient’s vitals are as follows:

~~~
Temperature: 99.8°F
Heart Rate: 105 bpm
Blood Pressure: 140/90 mmHg
Respiratory Rate: 18 breaths/min
Oxygen Saturation: 98% on room air
~~~

Please specify what aspects of the patient’s medical history, surgical history, and physical exam you would like to know.

USER: I would like to know what medications he is taking, what past surgeries he has had, and how he looks and where he is tender in his abdomen - is he guarding / does he have a history of Crohn’s disease / UC / has he ever had diverticulitis?

BOT: The patient’s medical history is positive for type 2 diabetes mellitus and hypertension, for which he is taking Metformin and Lisinopril respectively. He denies a history of Crohn’s disease, ulcerative colitis, or previous episodes of diverticulitis. He has no known drug allergies. His surgical history includes a cholecystectomy 10 years ago due to gallstones. On physical examination, the patient appears in moderate distress due to pain. His abdomen is soft, but there is tenderness and guarding in the left lower quadrant. No palpable masses or hernias are noted. His bowel sounds are slightly decreased. No other significant findings are noted on his physical examination.

USER: OK thank you. When was the last time he ate. Is he having any nausea? Is he a smoker? Any allergies to medications? Any history of bleeding or clotting disorders? Any history of cancer in the family?

BOT: The patient reports his last meal was approximately 12 hours ago. He denies any current feelings of nausea or vomiting. He quit smoking 15 years ago but had a 20 pack-year history prior to quitting. He reports no known drug allergies. He denies any personal history of bleeding or clotting disorders. Regarding his family history, his father had colorectal cancer and his mother had breast cancer.

USER: Any recent weight loss? Has he had a colonoscopy? And if so what did they find? I would ask the ER to draw labs, including a CBC and BMP and review the CT imaging that they would have already ordered before they called me.

BOT: The patient denies any recent unintentional weight loss. His last colonoscopy was performed 5 years ago and was normal, with no polyps or masses identified. Here are the lab results:

~~~
White Blood Cell Count: 13,000 per microliter
Hemoglobin: 14.6 g/dL
Platelet Count: 250,000 per microliter
Sodium: 138 mEq/L
Potassium: 4.5 mEq/L
Chloride: 100 mEq/L
Bicarbonate: 24 mEq/L
Blood Urea Nitrogen: 20 mg/dL
Creatinine: 1.0 mg/dL
Glucose: 180 mg/dL
~~~

The CT scan of the abdomen and pelvis reveals multiple diverticula in the sigmoid colon, one of which shows surrounding fat stranding and a small amount of free air. There is no evidence of a large abscess or mass.

USER: Thank you - this sounds like a Hinchey class II vs III acute diverticulitis, and it sounds like this is the patient’s first episode. I would make sure the patient is on IVF for fluid resuscitation, and make sure that he responds appropriately to fluid, and start IV antibiotics Zosyn. I would counsel the patient that if he doesn’t get better he may need an operation, particularly given the CT findings which have some worrisome features. I would counsel him on what non-response to non-operative management looks like as well as discuss the steps of potential surgical intervention, including a Hartman’s procedure. Can you tell me how he does with the fluid running and IV antibiotics?

BOT: The patient tolerates the IV fluids well and his heart rate reduces to 90 bpm. His pain seems to be better controlled with IV analgesics and he reports a significant decrease in pain with the initiation of the antibiotics.

However, after 48 hours of conservative management, the patient develops a fever of 101.5°F and increased abdominal pain. His white blood cell count has increased to 15,000 per microliter. His heart rate is 110 bpm and his blood pressure is stable at 135/85 mmHg. On examination, his abdomen is more distended and there is increased tenderness in the left lower quadrant. What would you like to do next?

USER: This patient has unfortunately failed non-operative management and needs an operation emergently; I would make sure the patient is NPO, place a NG tube and would call the OR and get him ready to do an exploratory laparotomy with plan for Hartmann’s procedure, which is where we take out his diseased colon and a portion of the diseased rectum, and leave an end colostomy. I would get a type and screen. I would also counsel him that he could get sicker afterwards and will likely go to the ICU and that he would need a bag colostomy for 3 to 6 months while he recovers and his tissue inflammation subsides, allowing for a takedown procedure. I would also call the stoma nurses to have him marked appropriately for an ostomy.

BOT: The patient is prepared for surgery as per your instructions. He understands the procedure and the potential need for ICU admission post-operatively. He is consented for an exploratory laparotomy with a planned Hartmann’s procedure. Can you please outline the steps for the Hartmann’s procedure?

USER: Yes, first I would do an exploratory laparotomy with a midline incision extending subxiphoid to the infraumbilical, to expose the abdomen and allow exploration of the abdomen, including the small and large intestine. I would first focus on examining the sigmoid colon and identifying the perforation. I would then wash out any purulence. I would next mobilize the sigmoid colon from the pelvic promontory extending up along the White line of Toldt to the splenic flexure, taking care to stay in the same plane and ensure that the ureter is not damaged, then ensure I have sufficient length to bring up an end colostomy. I would staple off the proximal colon and include a red rubber to decompress the colon, suctioning it, I would then staple off the distal end of the rectosigmoid to create a rectal pouch where there is healthy viable tissue, then I would create a circular opening with a cruciate incision through the fascia and bring up the proximal colon that has been stapled off as my end colostomy, then brooke the colon to create a colostomy with 3-0 and 4-0 vicryl suture. I would then irrigate the abdomen in all 4 quadrants thoroughly with 4 L of saline and close the abdomen with fascial and skin layers.

BOT: Thank you. The surgery proceeds as planned without any intraoperative complications. The patient is transferred to the ICU postoperatively.

On postoperative day 2, the patient develops shortness of breath and his oxygen saturation drops to 90% on room air. His heart rate is 110 bpm, blood pressure is 135/85 mmHg, and respiratory rate is 22 breaths/min. What would you like to do next?

USER: I would send stat labs on the patient, along with a stat CXR, EKG, and troponins. I would also assess him at the bedside, put him on supplemental oxygen, and ensure his pain is controlled. I would also send a D-dimer and consider a CT PE protocol. I am concerned for a pulmonary embolism, even though the patient is on prophylactic anticoagulation.

BOT: The patient’s labs are as follows:

~~~
White Blood Cell Count: 12,000 per microliter
Hemoglobin: 11.0 g/dL
Platelet Count: 230,000 per microliter
Sodium: 138 mEq/L
Potassium: 4.0 mEq/L
Chloride: 100 mEq/L
Bicarbonate: 24 mEq/L
Blood Urea Nitrogen: 25 mg/dL
Creatinine: 1.3 mg/dL
Glucose: 200 mg/dL
Troponin: 0.01 ng/mL
D-dimer: 500 ng/mL FEU
~~~

The chest X-ray shows no evidence of pneumonia or pneumothorax but there is a small left-sided pleural effusion. The EKG shows sinus tachycardia with no ST segment changes.

Given the clinical picture and elevated D-dimer, a CT angiogram of the chest is performed, which reveals a segmental pulmonary embolism in the right lower lobe. What would you like to do next?

USER: I would start the patient on therapeutic heparin drip with a bolus dose and assess his response, based on PTT goals, as the patient has a developing postoperative AKI and would not be amenable to therapeutic Lovenox. I would consult my colleagues in Medicine and Hematology to also assess for hypercoagulability although he has many risk factors for a PE.

BOT: The patient is started on a heparin drip as per your instructions and his PTT is monitored closely. His oxygenation improves with the increased anticoagulation, and his AKI does not worsen. The medicine and hematology teams agree with your management plan and do not feel further hypercoagulability workup is indicated at this time. The patient’s respiratory status gradually improves over the next few days and by postoperative day 7, he is transferred to the step-down unit. Looking back, how do you feel about your performance?

USER: I would say average. How would you assess my performance and what steps would you give me to improve along the way with each of the steps outlined?

BOT: Assessment of Performance:

- Organized Approach and Solid Rationale: You approached the case systematically and your decision-making process was logical and evidence-based. You reacted appropriately to the patient’s changing status and escalated care as needed.
- Determination and Interpretation of Key Findings: You correctly identified the findings on CT scan and appropriately diagnosed the patient with acute diverticulitis. You also correctly identified the complication of pulmonary embolism and initiated appropriate management.
- Efficient Use of Clinical Knowledge: You demonstrated strong clinical knowledge in your diagnosis and management of acute diverticulitis and its complications.
- Error Avoidance: There were no major errors in your management.
- Recognition of Personal Limitations: You appropriately recognized when to escalate care and sought consultation from other specialties when needed.
- Flexibility in Patient’s Changing Course: You appropriately escalated the patient’s care when he did not respond to initial management.
- Surgical Judgment, Clinical Reasoning, and Problem-solving: You showed excellent surgical judgment in your decision to perform surgery and your plan for a Hartmann’s procedure. Your clinical reasoning was sound and you managed complications appropriately.

Areas for Improvement:

- In the initial evaluation, you could have asked for a urinalysis to rule out a urinary tract problem as a possible cause of the patient’s LLQ pain.
- While you did consider the possibility of a pulmonary embolism, earlier consideration and prophylaxis could have possibly prevented the patient’s postoperative PE.
- It would be beneficial to incorporate more frequent reassessment of the patient’s status during their ICU stay, especially when there are changes in their condition, such as new shortness of breath.

Education:

- Acute diverticulitis is characterized by the inflammation and infection of diverticula, which are outpouchings in the colonic wall. Patients typically present with left lower quadrant abdominal pain, fever, and an increased white blood cell count. Uncomplicated diverticulitis can often be managed with antibiotics, while complicated diverticulitis (e.g., abscess, perforation) may require surgical intervention. Complications can occur after any surgical procedure. In this case, the patient developed a postoperative pulmonary embolism, likely due to immobilization and the hypercoagulable state induced by surgery. It is critical to consider this diagnosis in any postoperative patient who develops unexplained shortness of breath or hypoxemia. Anticoagulation is the mainstay of treatment, with the choice of agent depending on renal function, bleeding risk, and other patient factors.

Question:

- A 65-year-old man presents with LLQ pain, fever, and an increased white blood cell count. CT reveals multiple diverticula in the sigmoid colon with surrounding fat stranding. What is the next best step in management?
  1. Oral antibiotics and outpatient follow-up
  2. Hospital admission and IV antibiotics
  3. Urgent surgical consultation for possible resection
  4. Placement of a percutaneous drainage catheter
- Answer: B

## Appendix C - Evaluation rubric utilized by reviewers

**Figure 3:**
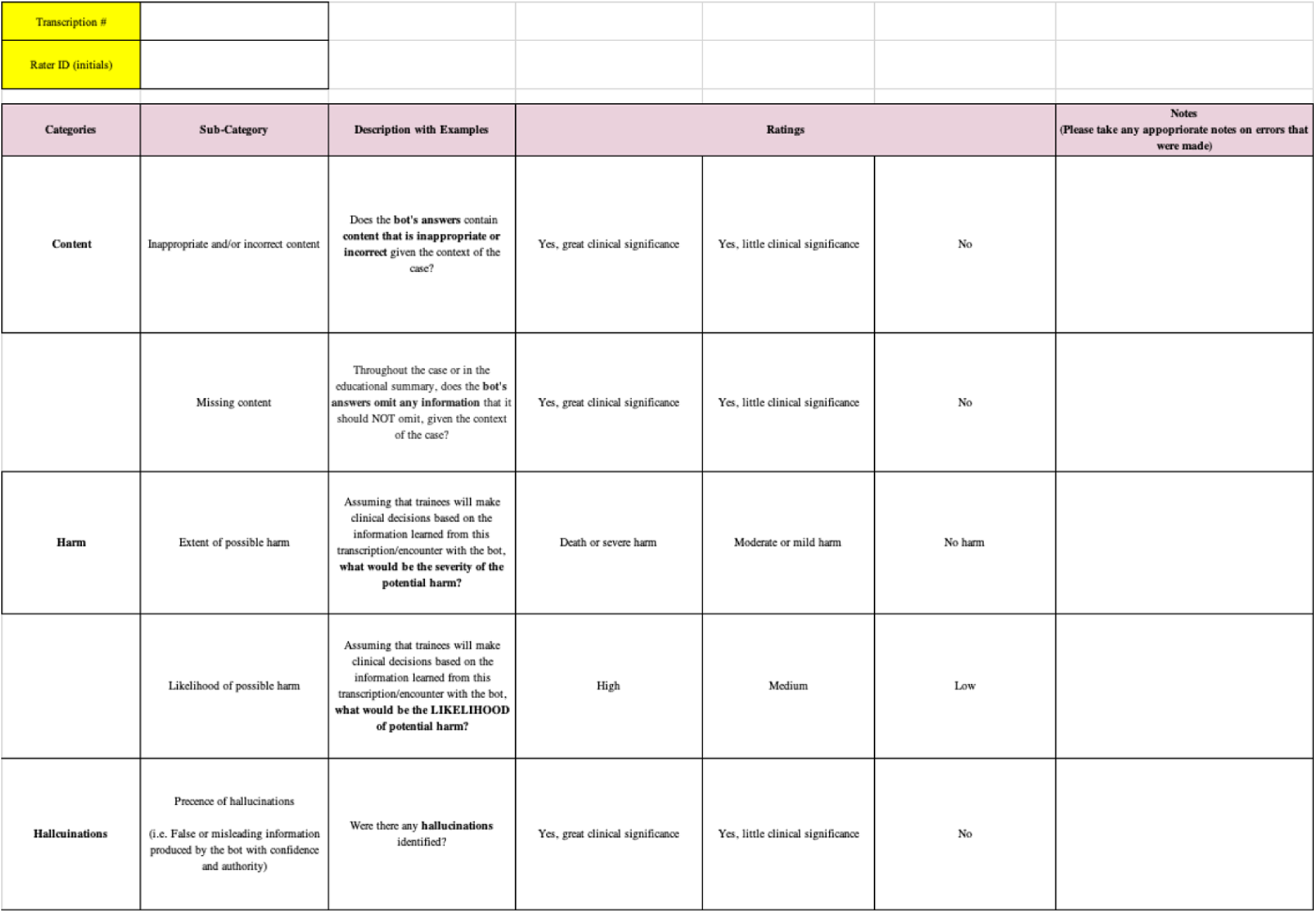
Evaluation rubric utilized by reviewers.

## References

[1] American board of surgery website. https://www.google.com/url?q=https://www.absurgery.org/get-certified/general-surgery/certifying-exam/&sa=D&source=docs&ust=1709940770349611&usg=AOvVaw0WwCVA3TkmONrbGQE4OYvf,. Accessed: 2024-5-01.

[2] Abbey L Fingeret, Tracey Arnell, John McNelis, Mindy Statter, Lisa Dresner, and Warren Widmann. Sequential participation in a Multi-Institutional mock oral examination is associated with improved american board of surgery certifying examination First-Time pass rate. J. Surg. Educ., 73(6):e95–e103, November 2016.

[3] Gary L Freed, Kelly M Dunham, and Dianne Singer. Use of board certification and recertification in hospital privileging: policies for general surgeons, surgical specialists, and nonsurgical subspecialists. Arch. Surg., 144(8):746–752, August 2009.

[4] Kelly M Dunham, Dianne Singer, and Gary L Freed. Use of board certification in ambulatory surgery center credentialing: a pilot study. J. Healthc. Manag., 54(1):31–42; discussion 42–3, 2009.

[5] Rachel O Reid, Mark W Friedberg, John L Adams, Elizabeth A McGlynn, and Ateev Mehrotra. Associations between physician characteristics and quality of care. Arch. Intern. Med., 170(16):1442–1449, September 2010.

[6] Jay B Prystowsky, Georges Bordage, and Joseph M Feinglass. Patient outcomes for segmental colon resection according to surgeon’s training, certification, and experience. Surgery, 132(4):663–70; discussion 670–2, October 2002.

[7] Daniel E Kendrick, Xilin Chen, Andrew T Jones, Michael Clark, Zhaohui Fan, Hoda Bandeh-Ahmadi, Greg Wnuk, Jason P Kopp, Beatriz Ibanez Moreno, John W Scott, Gurjit Sandhu, Jo Buyske, Justin B Dimick, and Brian C George. Is initial board certification associated with better early career surgical outcomes? Ann. Surg., 274(2): 220–226, August 2021.

[8] Timothy L Ruiz, Brandon Sellers, Aditya Devarakonda, Chase J Wehrle, and Tania K Arora. A novel mock oral curriculum for senior surgery residents: Results of a pilot study. J. Surg. Res., 277:92–99, September 2022.

[9] Taylor P Williams, Kevin J Hancock, V Suzanne Klimberg, Ravi S Radhakrishnan, Douglas S Tyler, and Alexander Perez. Learning to read: Successful Program-Based remediation using the surgical council on resident education (SCORE) curriculum. J. Am. Coll. Surg., 232(4):397–403, April 2021.

[10] Polina Zmijewski, Santh Prakash Lanka, Andrea Gillis, Brenessa Lindeman, Herbert Chen, and Jessica Fazendin. Regional mock oral board exercises for chief residents in general surgery. Am. J. Surg., 229:184–185, March 2024.

[11] Daniel Dante Yeh, John O Hwabejire, Ayesha Imam, John T Mullen, Douglas Smink, George Velmahos, and Marc DeMoya. A survey of study habits of general surgery residents. J. Surg. Educ., 70(1):15–23, 2013.

[12] Andrew T Jones, Thomas W Biester, Frank R Lewis, Jr, and Mark A Malangoni. Review courses for the american board of surgery certifying examination do not provide an advantage. Surgery, 158(4):890–6; discussion 896–8, October 2015.

[13] Edgar Guzman, Allen Babakhani, and Vijay K Maker. Improving outcomes on the ABS certifying examination: can monthly mock orals do it? J. Surg. Educ., 65(6):441–444, 2008.

[14] Daniel A London and Michael M Awad. The impact of an advanced certifying examination simulation program on the american board of surgery certifying examination passage rates. J. Am. Coll. Surg., 219(2):280–284, August 2014.

[15] Matthew D Cahn, Ace St John, and Stephen M Kavic. A scoping review of successful strategies for passing the american board of surgery certifying examination. Surg Open Sci, 17:12–22, January 2024.

[16] Walter E Longo and Amy L Friedman. Identifying programmatic deficiencies: the hidden value of the mock oral examination. Arch. Surg., 142(7):591–592, July 2007.

[17] Shari L Meyerson, Stuart Lipnick, and Edward Hollinger. The usage of mock oral examinations for program improvement. J. Surg. Educ., 74(6):946–951, May 2017.

[18] Gokulakkrishna Subhas, Stephen Yoo, Yeon-Jeen Chang, David Peiper, Mark J Frikker, David L Bouwman, Allen Silbergleit, Larry R Lloyd, and Vijay K Mittal. Benefits of mock oral examinations in a multi-institutional consortium for board certification in general surgery training. Am. Surg., 75(9):817–821, September 2009.

[19] Elise Paradis and Gary Sutkin. Beyond a good story: from hawthorne effect to reactivity in health professions education research. Med. Educ., 51(1):31–39, January 2017.

[20] Shawn Y Holmes. Mitigating the hawthorne effect using computer simulations. pages 175–187, 2011.

[21] A Karpathy. Intro to large language models. https://www.youtube.com/watch?v=zjkBMFhNj_g, November 2023. Accessed: 2023-11-30.

[22] Tejas S Sathe, Joshua Roshal, Ariana Naaseh, Joseph C L’Huillier, Sergio M Navarro, and Caitlin Silvestri. How I GPT it: Development of custom artificial intelligence (AI) chatbots for surgical education. J. Surg. Educ., 81(6):772–775, June 2024.

[23] Arya Rao, Michael Pang, John Kim, Meghana Kamineni, Winston Lie, Anoop K Prasad, Adam Landman, Keith J Dreyer, and Marc D Succi. Assessing the utility of ChatGPT throughout the entire clinical workflow. medRxiv, February 2023.

[24] Friederike Holderried, Christian Stegemann-Philipps, Lea Herschbach, Julia-Astrid Moldt, Andrew Nevins, Jan Griewatz, Martin Holderried, Anne Herrmann-Werner, Teresa Festl- Wietek, and Moritz Mahling. A generative pretrained transformer (GPT)-Powered chatbot as a simulated patient to practice history taking: Prospective, mixed methods study. JMIR Med Educ, 10:e53961, January 2024.

[25] Rajesh Bhayana, Satheesh Krishna, and Robert R Bleakney. Performance of ChatGPT on a radiology board-style examination: Insights into current strengths and limitations. Radiology, 307(5):e230582, June 2023.

[26] A Gilson, C Safranek, T Huang, V Socrates, L Chi, and others. How does ChatGPT perform on the medical licensing exams? the implications of large language models for medical education and knowledge assessment. MedRxiv, 2022.

[27] Tiffany H Kung, Morgan Cheatham, Arielle Medenilla, Czarina Sillos, Lorie De Leon, Camille Elepaño, Maria Madriaga, Rimel Aggabao, Giezel Diaz-Candido, James Maningo, and Victor Tseng. Performance of ChatGPT on USMLE: Potential for AI-assisted medical education using large language models. PLOS Digit Health, 2(2):e0000198, February 2023.

[28] Rohaid Ali, Oliver Y Tang, Ian D Connolly, Patricia L Zadnik Sullivan, John H Shin, Jared S Fridley, Wael F Asaad, Deus Cielo, Adetokunbo A Oyelese, Curtis E Doberstein, Ziya L Gokaslan, and Albert E Telfeian. Performance of ChatGPT and GPT-4 on neurosurgery written board examinations. Neurosurgery, 93(6):1353–1365, December 2023.

[29] Thomas Savage, Ashwin Nayak, Robert Gallo, Ekanath Rangan, and Jonathan H Chen. Diagnostic reasoning prompts reveal the potential for large language model interpretability in medicine. NPJ Digit Med, 7(1):20, January 2024.

[30] Zahir Kanjee, Byron Crowe, and Adam Rodman. Accuracy of a generative artificial intelligence model in a complex diagnostic challenge. JAMA, 330(1):78–80, July 2023.

[31] Eric Strong, Alicia DiGiammarino, Yingjie Weng, Andre Kumar, Poonam Hosamani, Jason Hom, and Jonathan H Chen. Chatbot vs medical student performance on Free-Response clinical reasoning examinations. JAMA Intern. Med., 183(9):1028–1030, September 2023.

[32] Banghao Chen, Zhaofeng Zhang, Nicolas Langreńe, and Shengxin Zhu. Unleashing the potential of prompt engineering in large language models: a comprehensive review. October 2023.

[33] Śebastien Bubeck, Varun Chadrasekaran, Ronen Eldan, Johannes Gehrke, Eric Horvitz, Ece Kamar, Peter Lee, Yin Tat Lee, Yuanzhi Li, Scott Lundberg, Harsha Nori, Hamid Palangi, Marco Tulio Ribeiro, and Yi Zhang. Sparks of Artificial General Intelligence: Early Experiments with GPT-4. arXIV, 2023.

[34] American board of surgery website. https://www.absurgery.org/get-certified/epas/general-surgery/,. Accessed: 2024-4-01.

[35] Ian M Kratzke, Sarah Lund, Amelia T Collings, Dominique L Doster, Julie M Clanahan, Andrea J H Williamson, Rachel M Jensen, Angela E Thelen, Amy Y Han, Rebecca S Gates, and Ladonna E Kearse. A novel approach for the advancement of surgical education: the collaboration of surgical education fellows (CoSEF). Global Surgical Education - Journal of the Association for Surgical Education, 1(1):38, September 2022.

[36] Karan Singhal, Shekoofeh Azizi, Tao Tu, S Sara Mahdavi, Jason Wei, Hyung Won Chung, Nathan Scales, Ajay Tanwani, Heather Cole-Lewis, Stephen Pfohl, Perry Payne, Martin Seneviratne, Paul Gamble, Chris Kelly, Abubakr Babiker, Nathanael Schärli, Aakanksha Chowdhery, Philip Mansfield, Dina Demner-Fushman, Blaise Agüera Y Arcas, Dale Webster, Greg S Corrado, Yossi Matias, Katherine Chou, Juraj Gottweis, Nenad Tomasev, Yun Liu, Alvin Rajkomar, Joelle Barral, Christopher Semturs, Alan Karthikesalingam, and Vivek Natarajan. Large language models encode clinical knowledge. Nature, 620 (7972):172–180, August 2023.

[37] Tamara Williams, Marilyn Szekendi, Stephen Pavkovic, Wanda Clevenger, and Julie Cerese. The reliability of AHRQ common format harm scales in rating patient safety events. J. Patient Saf., 11(1):52–59, March 2015.

[38] Ziwei Ji, Nayeon Lee, Rita Frieske, Tiezheng Yu, Dan Su, Yan Xu, Etsuko Ishii, Ye Jin Bang, Andrea Madotto, and Pascale Fung. Survey of hallucination in natural language generation. ACM Comput. Surv., 55(12):1–38, March 2023.

[39] Jacob Cohen. A coefficient of agreement for nominal scales. Educ. Psychol. Meas., 20 (1):37–46, April 1960.

[40] Mary L McHugh. Interrater reliability: the kappa statistic. Biochem. Med., 22(3): 276–282, 2012.

[41] Ishith Seth, Bryan Lim, Jevan Cevik, Foti Sofiadellis, Richard J Ross, Roberto Cuomo, and Warren M Rozen. Utilizing GPT-4 and generative artificial intelligence platforms for surgical education: an experimental study on skin ulcers. Eur. J. Plast. Surg., 47(1), January 2024.

[42] Gunther Eysenbach. The role of ChatGPT, generative language models, and artificial intelligence in medical education: A conversation with ChatGPT and a call for papers. JMIR Med Educ, 9:e46885, March 2023.

[43] Yunzhe Xue, Andrew Hu, Rohit Muralidhar, Justin W Ady, Advaith Bongu, and Usman Roshan. An AI system for evaluating pass fail in fundamentals of laparoscopic surgery from live video in realtime with performative feedback. In 2023 IEEE International Conference on Bioinformatics and Biomedicine (BIBM), pages 4167–4171. IEEE, December 2023.

[44] Kai Siang Chan and Nabil Zary. Applications and challenges of implementing artificial intelligence in medical education: Integrative review. JMIR Med Educ, 5(1):e13930, June 2019.

[45] Mert Karabacak, Burak Berksu Ozkara, Konstantinos Margetis, Max Wintermark, and Sotirios Bisdas. The advent of generative language models in medical education. JMIR Med Educ, 9:e48163, June 2023.

[46] Monika Hooda, Chhavi Rana, Omdev Dahiya, Ali Rizwan, and Md Shamim Hossain. Artificial intelligence for assessment and feedback to enhance student success in higher education. Math. Probl. Eng., 2022, May 2022.

[47] Riley Scherr, Faris F Halaseh, Aidin Spina, Saman Andalib, and Ronald Rivera. ChatGPT interactive medical simulations for early clinical education: Case study. JMIR Med Educ, 9:e49877, November 2023.

[48] Moloud Abdar, Farhad Pourpanah, Sadiq Hussain, Dana Rezazadegan, Li Liu, Mohammad Ghavamzadeh, Paul Fieguth, Xiaochun Cao, Abbas Khosravi, U Rajendra Acharya, Vladimir Makarenkov, and Saeid Nahavandi. A review of uncertainty quantification in deep learning: Techniques, applications and challenges. Inf. Fusion, 76:243–297, December 2021.

[49] Jakob Gawlikowski, Cedrique Rovile Njieutcheu Tassi, Mohsin Ali, Jongseok Lee, Matthias Humt, Jianxiang Feng, Anna Kruspe, Rudolph Triebel, Peter Jung, Ribana Roscher, Muhammad Shahzad, Wen Yang, Richard Bamler, and Xiao Xiang Zhu. A survey of uncertainty in deep neural networks. Artificial Intelligence Review, 56(1): 1513–1589, October 2023.

[50] Harsha Nori, Yin Tat Lee, Sheng Zhang, Dean Carignan, Richard Edgar, Nicolo Fusi, Nicholas King, Jonathan Larson, Yuanzhi Li, Weishung Liu, Renqian Luo, Scott Mayer McKinney, Robert Osazuwa Ness, Hoifung Poon, Tao Qin, Naoto Usuyama, Chris White, and Eric Horvitz. Can generalist foundation models outcompete Special-Purpose tuning? case study in medicine. November 2023.

[51] T Gao. Prompting: Better ways of using language models for NLP tasks. https://thegradient.pub/prompting/, 2021. Accessed: 2024-4-NA.

[52] Angels Balaguer, Vinamra Benara, Renato Luiz de Freitas Cunha, Roberto de M. Estevão Filho, Todd Hendry, Daniel Holstein, Jennifer Marsman, Nick Mecklenburg, Sara Malvar, Leonardo O Nunes, Rafael Padilha, Morris Sharp, Bruno Silva, Swati Sharma, Vijay Aski, and Ranveer Chandra. RAG vs fine-tuning: Pipelines, tradeoffs, and a case study on agriculture, 2024.

[53] P Bhavsar. RAG vs fine-tuning vs both: a guide for optimizing LLM performance. https://www.rungalileo.io/blog/optimizing-llm-performance-rag-vs-finetune-vs-both, 2023. Accessed: 2024-3-NA.

[54] Richard H Bell, Jr, Thomas W Biester, Arnold Tabuenca, Robert S Rhodes, Joseph B Cofer, L D Britt, and Frank R Lewis, Jr. Operative experience of residents in US general surgery programs: a gap between expectation and experience. Ann. Surg., 249 (5):719–724, May 2009.

[55] Robert D Acton, Jeffrey G Chipman, Michelle Lunden, and Connie C Schmitz. Unanticipated teaching demands rise with simulation training: strategies for managing faculty workload. J. Surg. Educ., 72(3):522–529, 2015.

[56] Benjamin Zendejas, Amy T Wang, Ryan Brydges, Stanley J Hamstra, and David A Cook. Cost: the missing outcome in simulation-based medical education research: a systematic review. Surgery, 153(2):160–176, February 2013.

[57] J Ker, G Hogg, and N Maran. Cost-effective simulation. Cost effectiveness in medical education, pages 61–71, March 2021.

[58] Taojun Hu and Xiao-Hua Zhou. Unveiling LLM evaluation focused on metrics: Challenges and solutions, 2024.

[59] An Yang, Kai Liu, Jing Liu, Yajuan Lyu, and Sujian Li. Adaptations of ROUGE and BLEU to better evaluate machine reading comprehension task, 2018.

